# Chaos, Percolation and the Coronavirus Spread: the Italian case

**DOI:** 10.1101/2020.04.10.20060616

**Authors:** Aldo Bonasera, G. Bonasera, Suyalatu Zhang

## Abstract

A model based on chaotic maps and turbulent flows is applied to the spread of Coronavirus for each Italian region in order to obtain useful information and help to contrast it. We divide the regions into different risk categories and discuss anomalies. The worst cases are confined between the Appenine and the Alps mountain ranges but the situation seem to improve closer to the sea. The Veneto region gave the most efficient response so far and some of their resources could be diverted to other regions, in particular more tests to the Lombardia, Liguria, Piemonte, Marche and V. Aosta regions, which seem to be worst affected. We noticed worrying anomalies in the Lazio, Campania and Sicilia regions to be monitored. We stress that the number of fatalities we predicted on March 12 has been confirmed daily by the bulletins. This suggests a change of strategy in order to reduce such number maybe moving the weaker population (and negative to the virus test) to beach resorts, which should be empty presently. The ratio deceased/positives on April 4, 2020 is 5.4% worldwide, 12.3% in Italy, 1.4% in Germany, 2.7% in the USA, 10.3% in the UK and 4.1% in China. These large fluctuations should be investigated starting from the Italian regions, which show similar large fluctuations.

## Introduction

The 2020 widespread of the Coronavirus or COVID-19 virus could be compared to the spread of the Red Weevil (Rhynchophorus Ferrugineus) in the Mediterranean or fires in California. They start in one or more localized places and quickly spread over larger and larger regions until it becomes difficult to stop them. After that the spread continues to ‘affect’ more and more regions until there is some ‘fuel’, i.e. palm trees for the Red Weevil or woods for the fire. This mechanism is similar to physical systems, for instance turbulent flow or chaotic maps^1-6^, where a small perturbation grows exponentially and then saturates to a finite value. These at first sight different systems have some common features: a small perturbation, which we will indicate as d_0_, grows exponentially with a coefficient *γ*, the Lyapunov exponent, and finally saturates^1-3^ to a value *d*_∞_>>d_0_. The fact that every chaotic system saturates to a finite value, even though might be very large, indicates that the ‘phase-space’ is however limited and reflects some conservation laws, such as energy conservation for a physical system or the number of palm trees for the Red Weevil. We can write the number of people for instance positives to the virus (or deceased for the same reason) as:

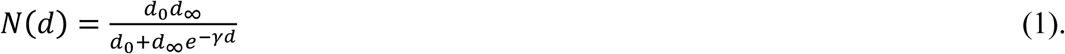

In the equation, d gives the time, in days, from the starting of the epidemic, or the time from the beginning of the tests to isolate the virus. At time d=0, N(0)=d_0_ which is the very small value (or group of people) from which the infection started. In the opposite limit, *d* →∞, *N*(∞)=*d*_0_, the final number of affected people by the virus. In a recent paper^7^ we have analyzed the 2003 SARS and the COVID-19 viruses using the equation written above and fitting the three parameters to the data. The model reproduces the data very well on a daily basis starting from March 12 for the Italy case^7^. This might be coincidental but it is further supported by the analysis of the virus spread in other countries^7^.

The first important result that we pointed out is that to have information on the number of positive to the virus (or fatalities) is not statistically relevant if we do not know the total number of tests performed each day and possibly the method chosen to perform the tests. The method to choose the people to be tested might be biased because of the large number of people affected and the limited amount of tests and facilities. We have been able to obtain quickly the total number of daily tests (and other relevant quantities) from https://github.com/pcm-dpc/COVID-19.

In the figure 1, we plot the total number of tests performed in all Italian regions starting from February 24,2020. The different regions are indicated by different colors and/or symbols. The reason for each color will be discussed more in detail later and we have divided them into the dark red, red, blue, cyan and green colors depending on the probability to find a positive to the virus in that region. So dark red gives the highest probability while green is the lowest. Notice that the Lombardy, Veneto and E. Romagna regions performed most test. The other important feature to notice is the change of slope after day 10 (starting from February 24,2020), which means that after that day the number of tests performed daily more than doubled. This change on the number of tests and the different number for different countries or regions make it difficult to make predictions on absolute values such as the total number of positives to the virus or any other quantities. Thus it is statistically more relevant to define probabilities for instance from the ratio of positives divided by the total number of tests or the number of fatalities divided by the total number of tests etc.

**Figure 1.**
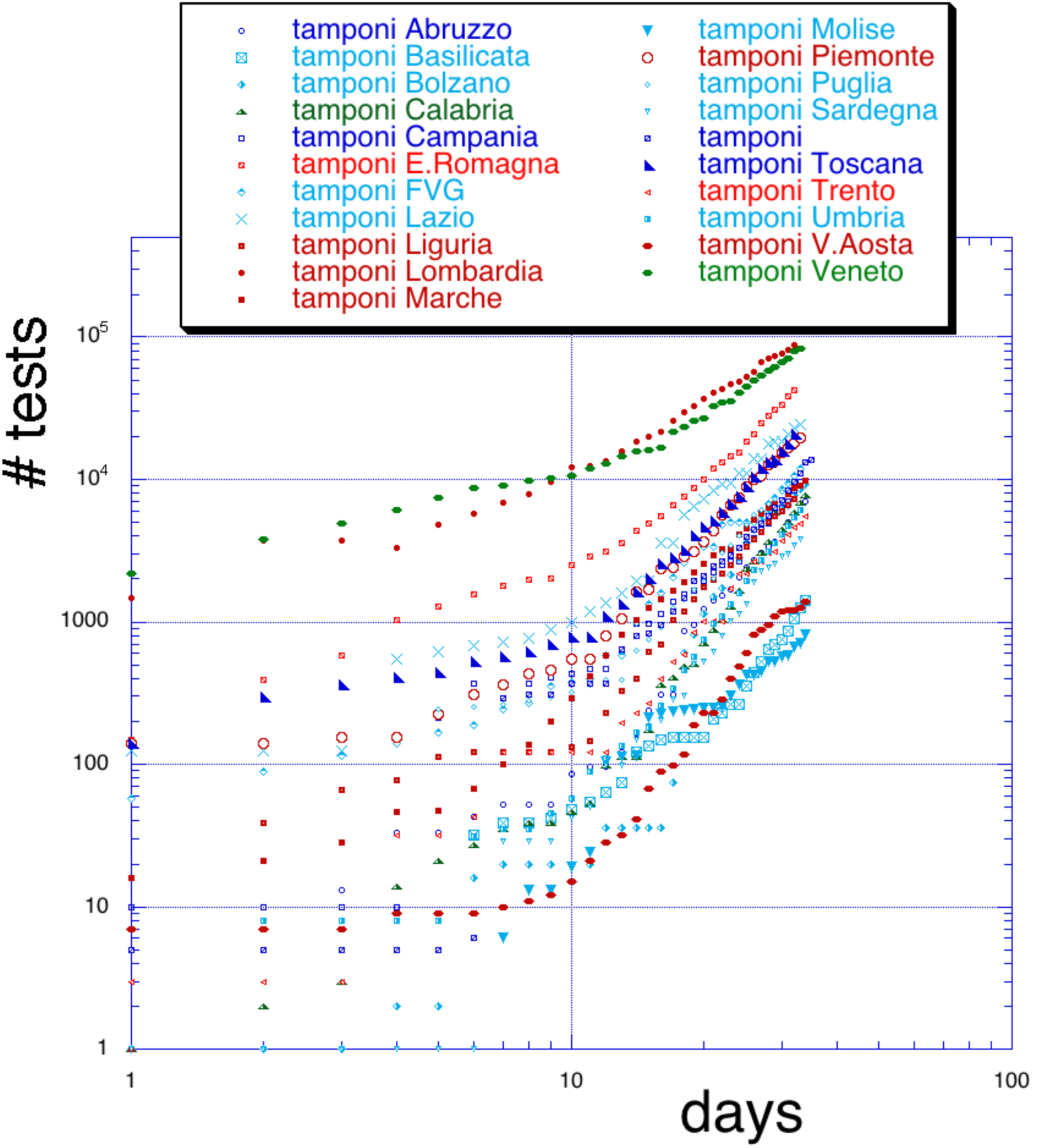
Number of tests as function of the day, starting from February 24, 2020. The different Italian regions are indicated in the inset.

The equation (1) is well suited to predict these probabilities once a fit has been performed on some preliminary results, we apply it first to Italy as a whole. In the figure 2, we show the results of the fit by fixing the parameters on March 12, 2020. A previous fit after the start of the epidemic was performed on the positives, it is given by the upper (black) points in the figure. As we saw in figure 1 after about 10 days the number of tests was more than doubled and also quarantine measures were taken by the Italian government. This led to the new fit on March 12 given by the cyan points in the figure. Since then we have not modified the fit but just added the new daily points which seem to follow the new fit for the positives up to April 4, 2020. As we can see there seem to be a decrease for the last points. A decrease from the prediction means that the probability to be positive to the virus is decreasing and social distancing plus other measures are giving some results. If we now analyze the fatalities, we first of all notice that the original prediction is followed by the data and we did not need to perform a new fit on March 12 at variance with the number of positives. Furthermore, the probability for fatalities seem to follow the prediction and little or no decrease is observed. Another important quantity that we plot is the ratio deceased/positives, which should be somewhat independent on the total number of tests if the method to choose the people to be tested does not change. As we can see the ratio keeps increasing and becomes larger than 10%. This is much higher than Germany for instance, which has a large number of positives as well, and can be statistically compared to Italy. Other countries like Spain and UK show similar values for the ratio like Italy. These large fluctuations might be attributed to the different health facilities, ventilators, hospital overcrowding etc. It is important to coordinate the action in different countries to try to understand the reason for the discrepancies and save many lives.

**Figure 2.**
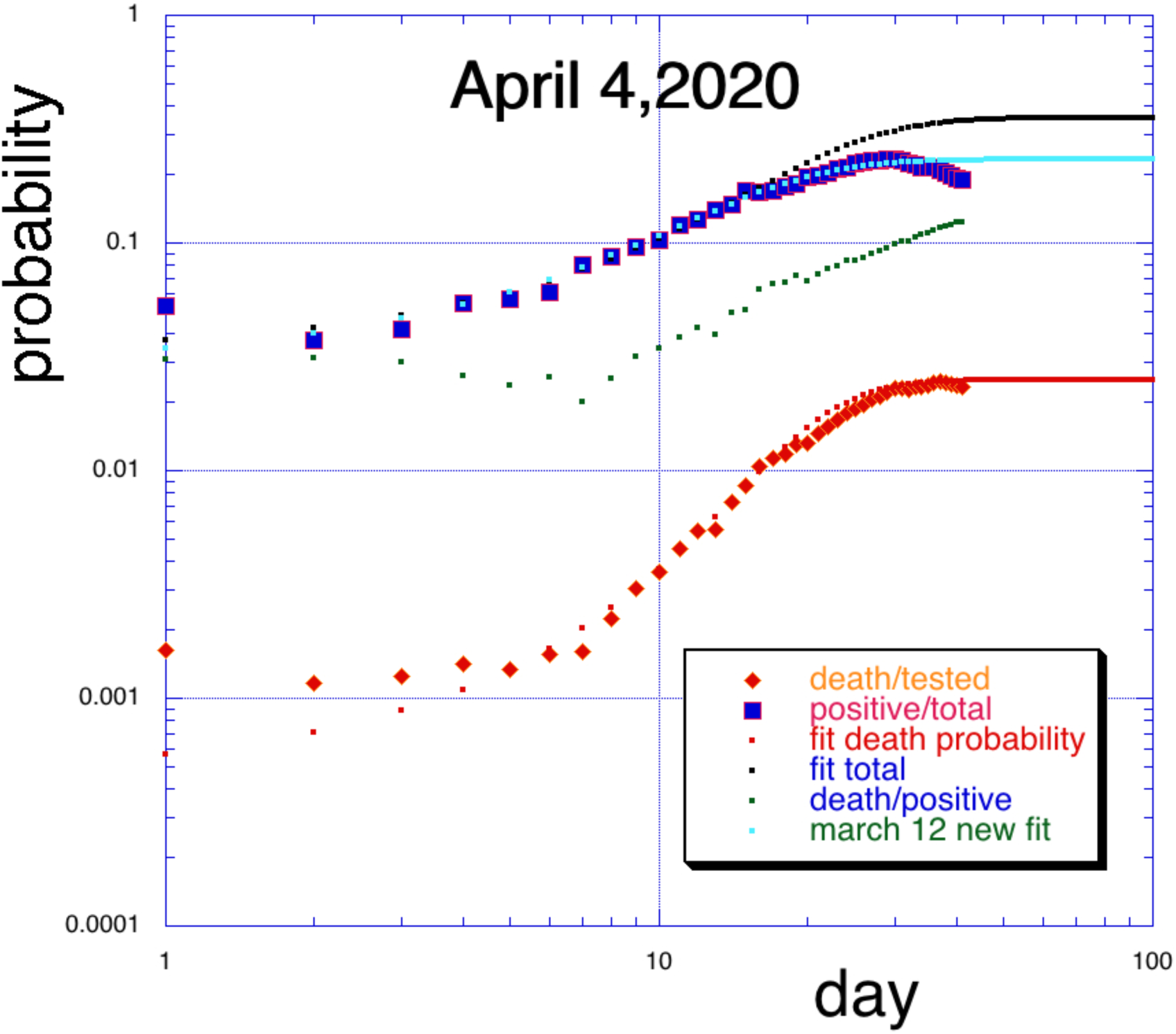
Italy: number of positives to the test (upper points) and number of fatalities (lower points) both normalized by the total number of tests performed (see figure 1) as function of the day from February 24, 2020. The green full squares in the middle give the ratio fatalities/positives and reaches above 10%. The smooth curves are the results of the fits using eq.(1). The fits were performed before March 12, 2020 while the actual data has been update daily to April 4, 2020.

One possibility for the large number of fatalities in Italy respect to other countries could be a time delay between being tested positive and passing away. In fact, in figure 2 we notice that the ratio is less than 3% before day 10, similar to other countries. However, China, Germany, Spain, UK and other countries have been contrasting the virus from more than 10 days already thus any transient effect should be finished. Another reason we could explore is hospital overcrowding which leads to a lack of resources to deal with the emergency. In figure 3 we plot the ratio=fatalities/positives as function of the number of people hospitalized each day. A clear correlation between the two variables is visible and we have parameterized it as:

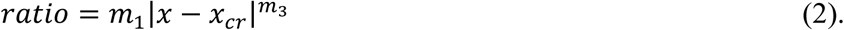

m_1_,x_cr_ and m_3_ are fitting parameters and are displayed in the figure. In particular x_cr_ gives a ‘critical’ value above which the ratio grows quickly. This parameterization is inspired by critical phenomena such as the liquid-gas (second order) phase transition in normal fluids and also in chaotic maps^1^. If taken literally this result would imply that hospitals should not admit more than 694±56 people daily, however as discussed above for the total number of tests, this plot would be more meaningful if we would know how many patients can the hospital accommodate in normal and safe conditions. In other words the number of hospitalized people might increase together with the number of hospitals involved thus to have an unequivocal correlation the total hospital capability should be known. The latter is not given in https://github.com/pcm-dpc/COVID-19 and, we hope, this will be addressed soon, together with the total number of ventilators available.

**Figure 3.**
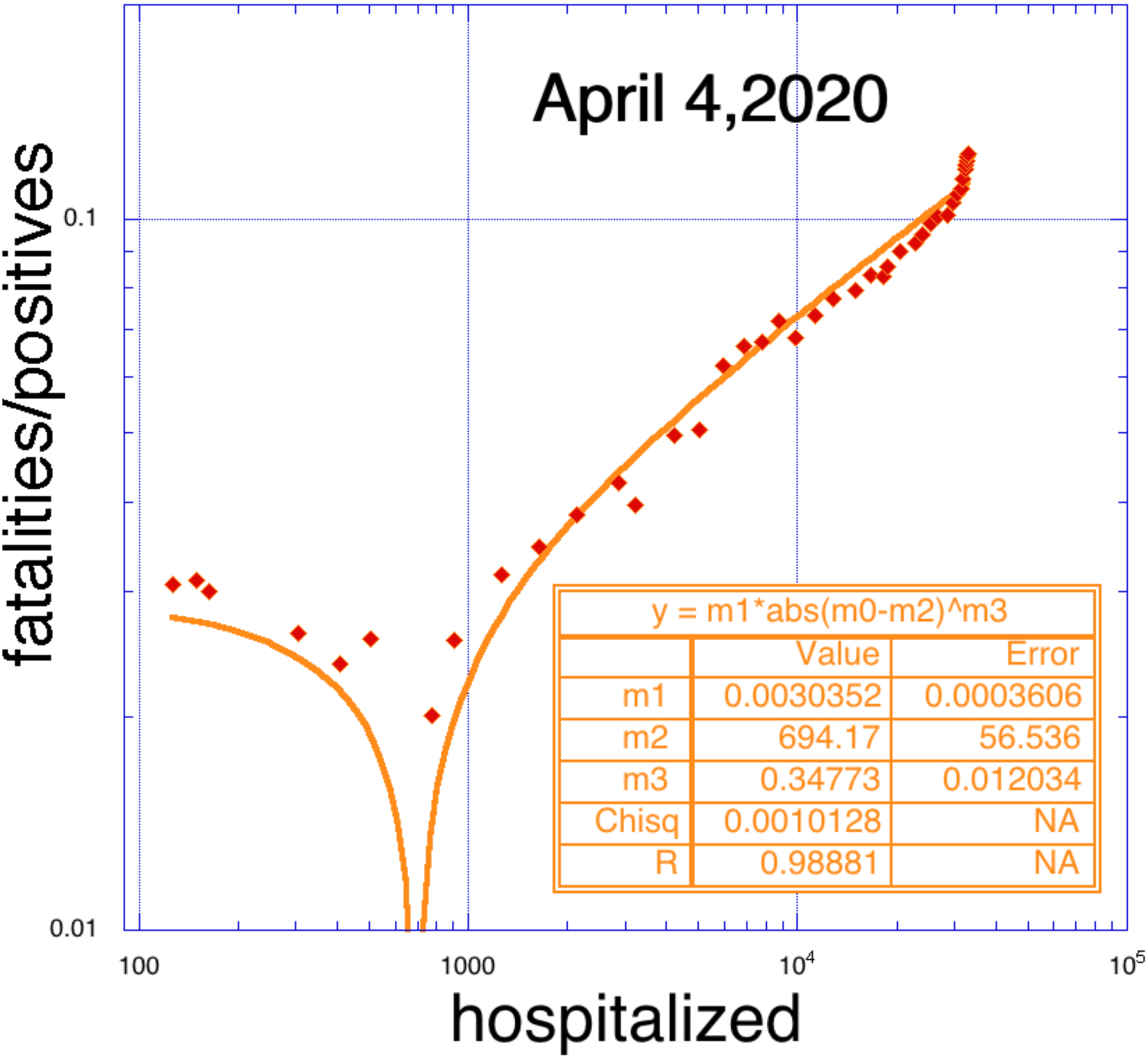
Ratio fatalities/positives as function of people hospitalized daily. The full line is the result of the fit given by eq.(2). The fit values are given in the inset.

In any case we expect that some kind of correlation like in figure 3 will remain. Other delocalized facilities should be organized to do a preliminary screening and admit to the hospital only the most severe cases, below the capability limits for each facility. Figure 2 shows that at most 1 person out of 5 tests positive to the virus but being admitted in an overcrowded hospital increases the probability of serious complications, i.e. more than 1 person out of 50 might die (in Germany it is roughly 1 out of 300 presently). An overcrowded hospital implies that the number of ventilators (which seem to be the last resource to fight the virus) is not sufficient.

Equation (1) gives a very good description of the probability of being infected by the virus but does not give us any hint on when the virus will stop its deadly action. We could in principle apply the same equation to the number of positives for instance without normalizing by the total number of tests. We have shown that this procedure might be meaningless if the number of tests performed daily changes or varies from region to region. However, from figure 1 we notice that after day 10 the number of test is a straight line, which implies that the number of tests per day is constant. Thus we may hope that equation (1) includes the trivial increase of daily tests in the 3 parameters and try to make predictions. This rather empirical method to predict the evolution of the spread might be justified only by the results. Once we get some confidence for some cases we can apply it to other cases keeping in mind that the total number of tests daily must be constant.

In the figure 4, we plot the total number of positives, dismissed healthy and fatalities as function of the day for the Italian case. As we can see the fit reproduces rather well the behavior at longer times and the data seem to saturate. For shorter times (below day 10) the model disagree with the data due to the fact that the number of tests performed daily increased. Thus the prediction of figure 4 should be taken with caution and we hope that at least the order of magnitude is correct. Refitting on April 5, 2020 increased the predictions to 1.43e5±1625 (1.35e5), 17904±296 (16162) and 28556±889 (20954) for positives, fatalities and dismissed respectively (in parentheses the values of the fit on March 31, 2020). The values of the fits performed at different times are slightly different which suggests that the fit is not convergent yet. However, the results are not much different which is a good sign together with the decrease observed in figure 2.

**Figure 4.**
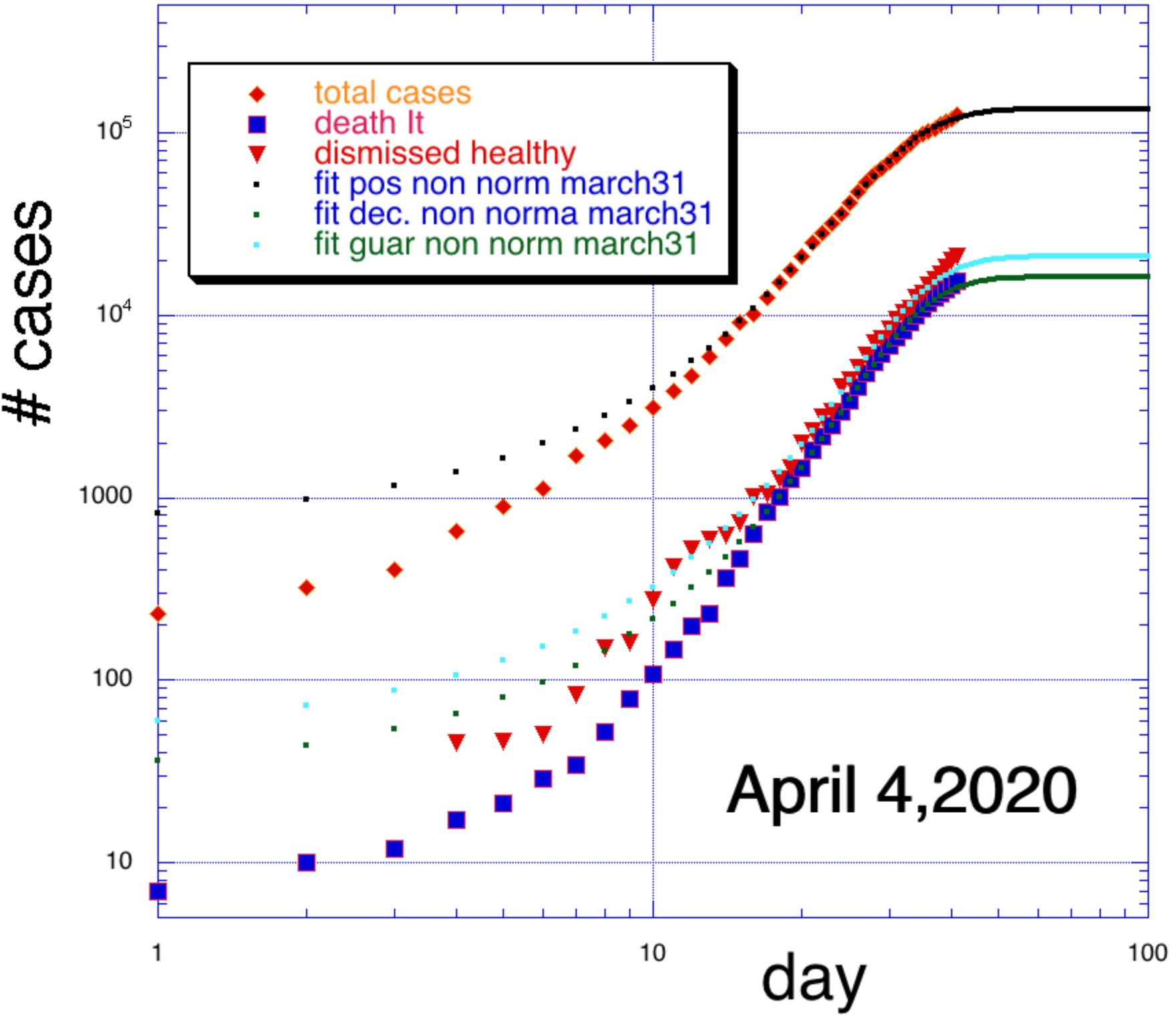
Total number of positives, dismissed healthy and fatalities as function of the day. The fits using equation (1) were performed on March 31 and new data have been added daily.

### Specializing the Model to the Lombardy region

We can specialize the previous results to each Italian region to get important information on the spreading and also to unveil anomalies, which could indicate new centers for the epidemic. We start with the Lombardy region, the most affected by the pandemic.

In figure 5 we plot the probabilities as function of the day similar to the Italy case reported in figure 2. The probability to be infected reaches almost 50% while the fatalities are up to about 7% and the ratio fatalities/positives is almost 20%, confirming, even if not needed, that Lombardy is the most affected region. Differently from figure 2 where a small decrease is observed at later days, Lombardy does not show any decrease but it seems very close to saturation. These results must be clarified since if this probability would reflect the actual population then 1 person out of 2 carries the virus. This is not the case since the method to choose the people to be tested is biased, i.e. those who show strong signs of infections are tested until there are no more tests available. This is one reason why some resources should be diverted temporarily to the Lombardy region starting for instance from the Veneto region, see figure 1.

**Figure 5.**
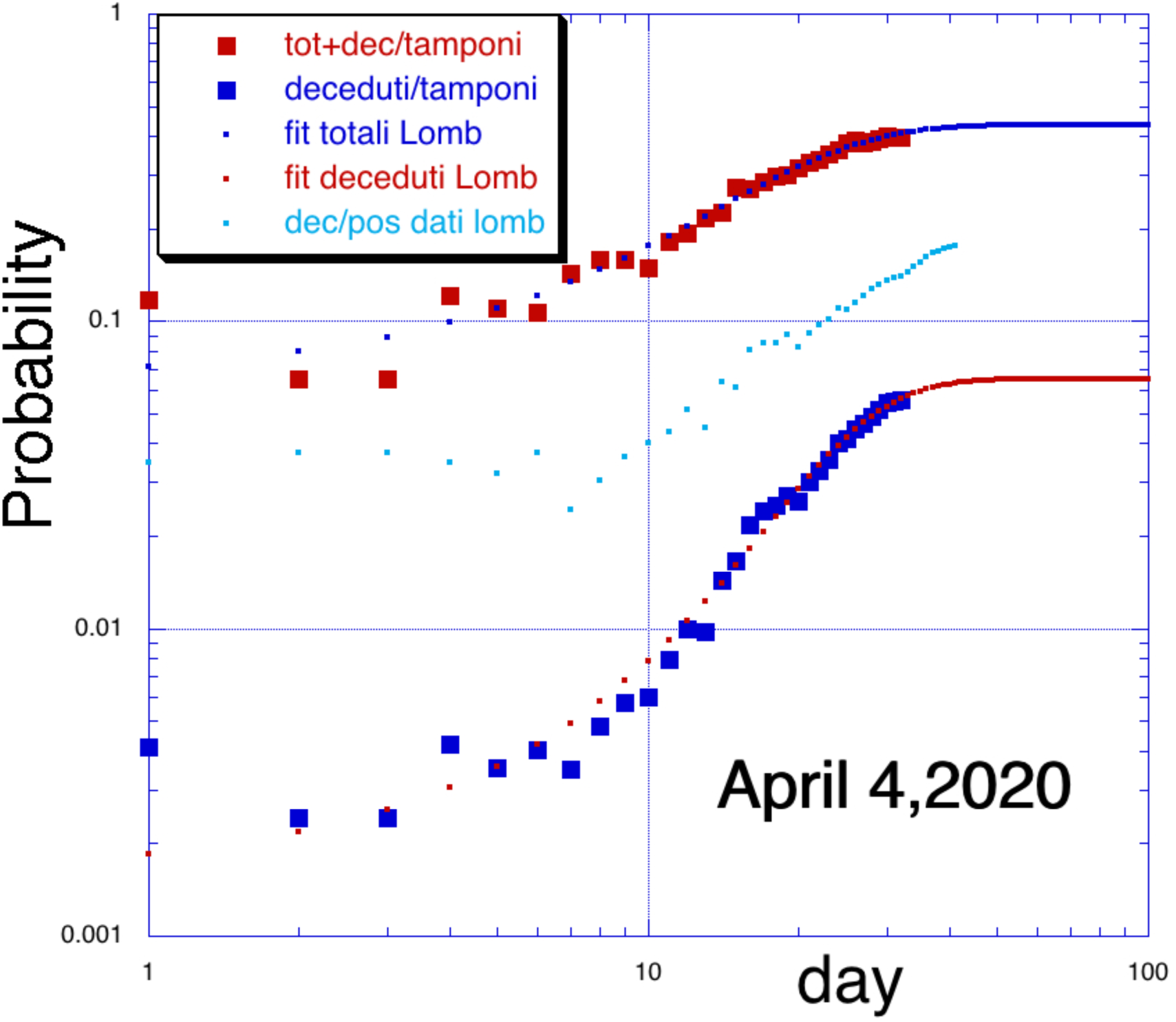
Same as figure 2, for the Lombardy region. The fits were performed on March 26 but the data has been update to April 4,2020.

The large number of fatalities may be due to hospital overcrowding and lack of equipment notably ventilators which seem to be the best tool to fight the infection or at least give more time to the organism to produce antibodies. Following the previous result for the Italian case, we plot in figure 6 the ratio=fatalities/positives as function of the number of people hospitalized each day for the Lombardy and the Veneto regions. The behavior and the fit indicated in the figure confirms that Lombardy is the region contributing mostly to the epidemics. The smaller ratio for the Veneto region is consistent with the lower number of hospitalized persons but higher than other nations like Germany.

**Figure 6.**
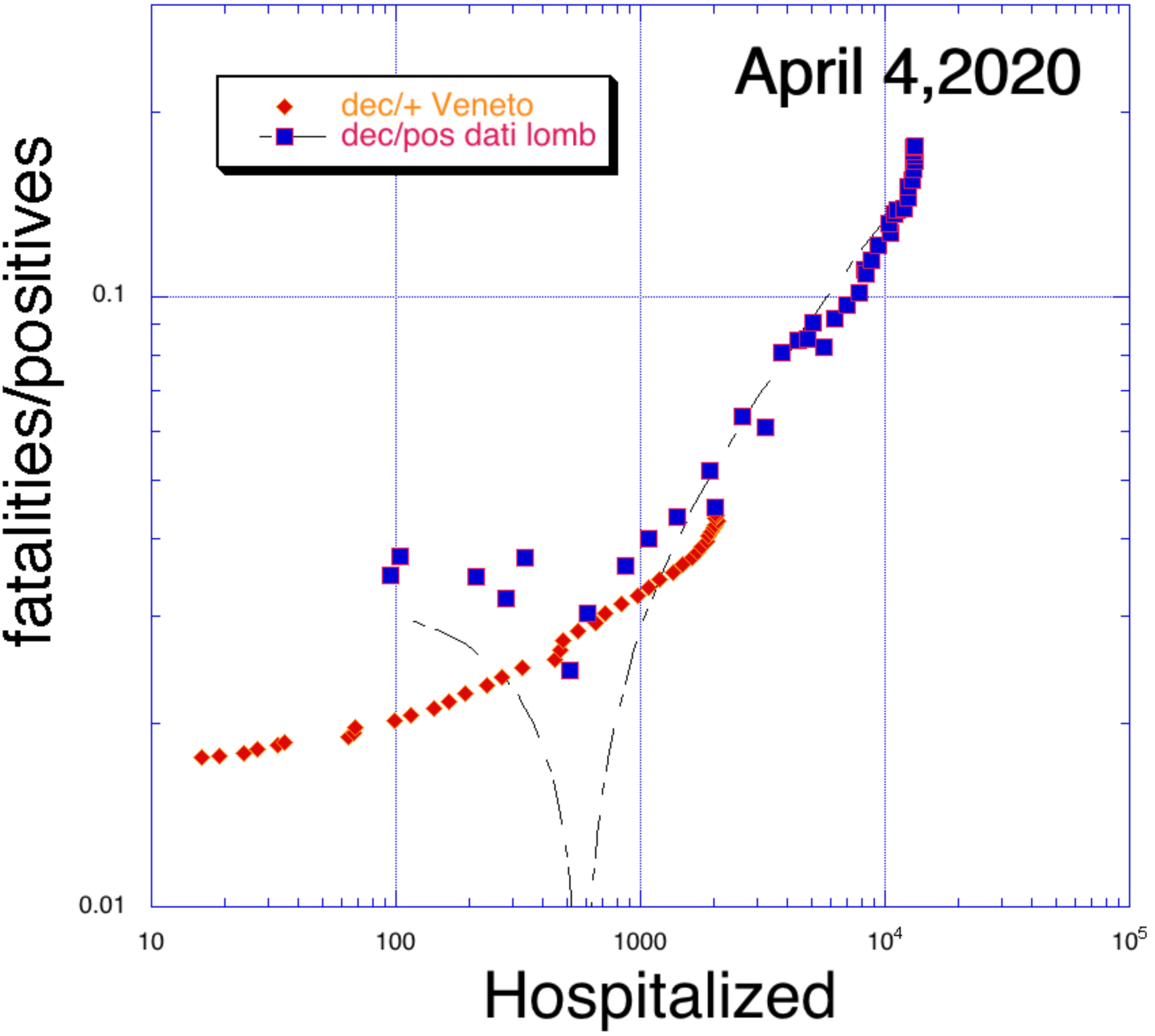
Same as figure 3, for the Lombardy and Veneto regions.

In figure 7, we use equation (1) to predict the total number of people affected by the virus. The fits on April 4,2020 gave 53286±596, 10310±174 and 15943±501 for positives, fatalities and dismissed cases respectively. These numbers can be compared to the National case given in the previous section: they contribute more than 50%. Again the fits are not so good at shorter times due to the changing number of tests performed daily.

**Figure 7.**
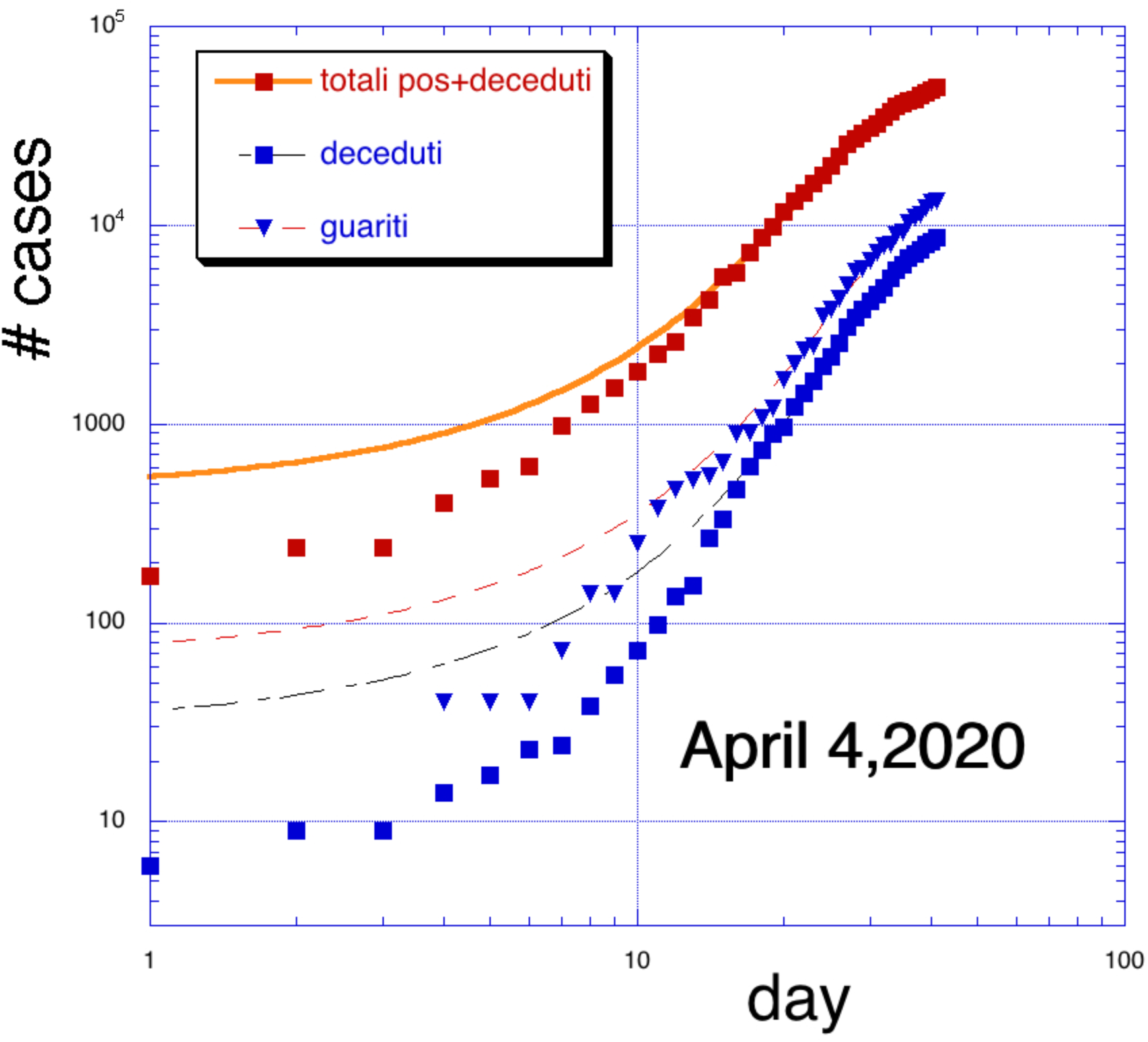
Same as figure 4, for the Lombardy region.

### All Regions

Following the methods outlined in the previous section, we can summarize the results for each region by looking in detail to all the fits.

In figure 8 we plot the probability of being tested positive as function of time for all the Italian regions. The fits were performed using equation 1 and following the method explained in the previous sections. All the fits were performed on March 31,2020. Different regions were grouped with different colors and symbols in the figure in order to distinguish them according to the probability. The regions with the highest probability are Lombardy, Liguria, Piemonte, Marche and V.Aosta. Some might come as a surprise but recall that we are plotting the positives divided by the total number of tests. The Veneto region, which is one of the most affected, is represented by the green color, one of the lowest probabilities. This is due to the high number of tests performed in that region as can be seen in figure 1. Comparing figures 1 and 8 one could find reasons to shift resources as needed. As a preliminary method, we should shift resources from regions, which have less than 15% probability (below the cyan color in the figure) to be infected to higher probability regions. This could be done on a temporary basis, say for a week to see if the probabilities, for the most affected regions, decrease. Of course, ideally to increase the total number of tests everywhere would be the best solution. A probability say of 30% means that almost 1 person out of 3 is affected, thus even in apartments with more than 3 persons living in it, social distancing and other precautions should be enforced. This virus might be asymptomatic, i.e. we might carry it and show no signs, thus the importance to obtain better estimates of the probabilities.

**Figure 8.**
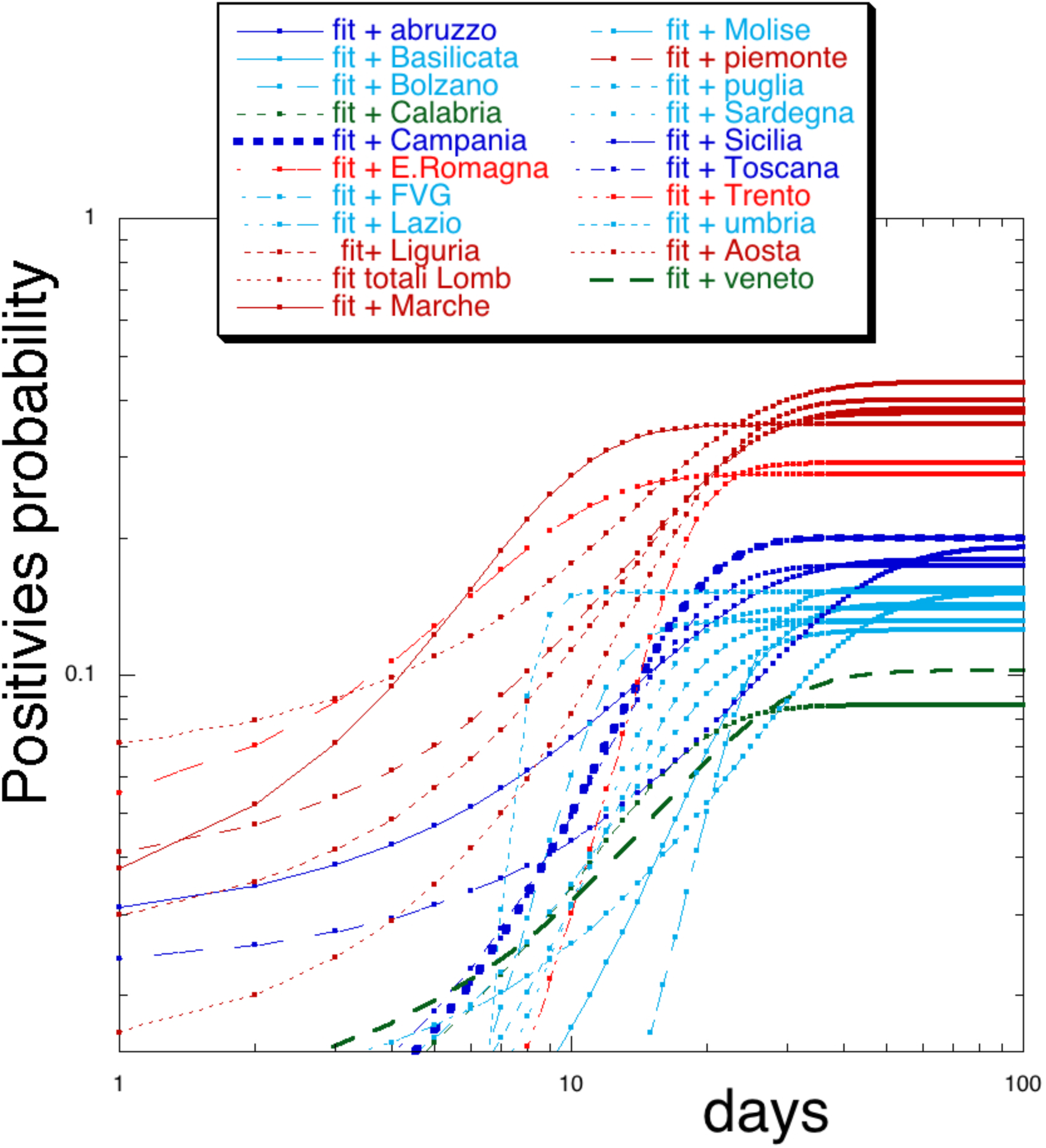
Probability of testing positive to the virus as function of time, from equation (1). Different regions are grouped by different colors.

It is also important to notice that some regions reach the saturation value earlier than others. For instance the Marche (dark red symbols) and E. Romagna (red symbols) regions saturate earlier than their respective color groups. A faster saturation might not be good because it might give not enough time to the hospitals to deal with large number of patients arriving at the same time. The Umbria region (cyan symbols) is the one that saturates first fortunately with a small number of positives and a large number of tests. For reference the same probability as in figure 8 for Germany is about 10%, https://www.worldometers.info/coronavirus/#countries, which would be the first goal: perform enough tests to be sure that all regions are effectively below 10% probability.

In figure 9 we plot the fatality probability as function of time. Notice that the color grouping is not maintained and in particular we notice the large ‘jump’ of the Lazio region (cyan symbols) which goes ‘three colors up’ while there is some ‘improvement’ for Piedmont (dark red symbols). The Veneto region, which is very close to the pandemic center (Lombardy), has a fatality rate less than 0.5%. This is the minimum goal, which can be reached by many other regions improving on the support system and maybe moving resources around. The reason(s) why the Lazio region is performing so poorly is not clear. The ‘best performing’ regions like Calabria, Basilicata and Umbria are located in the center, southern part of Italy, far away from the center of the infection^7^. Sicily is also a surprise being the most southern region and having such a large fatality rate, 2.2%. For comparison the probability for Germany is less than 0.2%.

**Figure 9.**
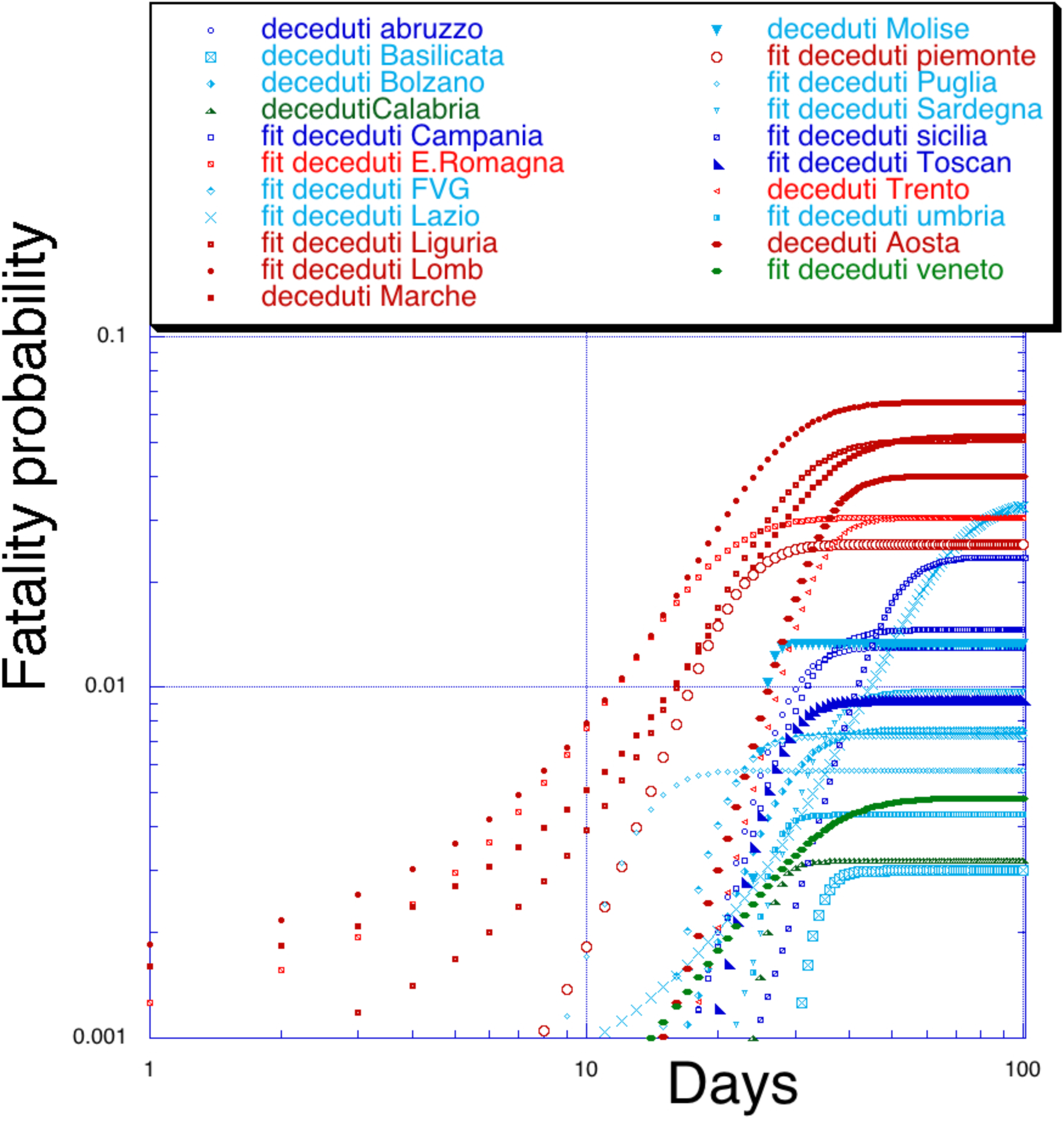
Fatalities (probability) as function of time for all the Italian regions. Notice the switch of color ordering for some regions compared to figure 8.

In figure 10 we plot the ratio=fatalities/positives, a quantity we have discussed the in previous sections. We mentioned the fact that such a ratio is less than 2% for Germany on April 5,2020 a value similar to the Basilicata region only, but while Germany had 100000 positives and 1600 deaths, Basilicata had 278 and 13 cases respectively.

**Figure 10.**
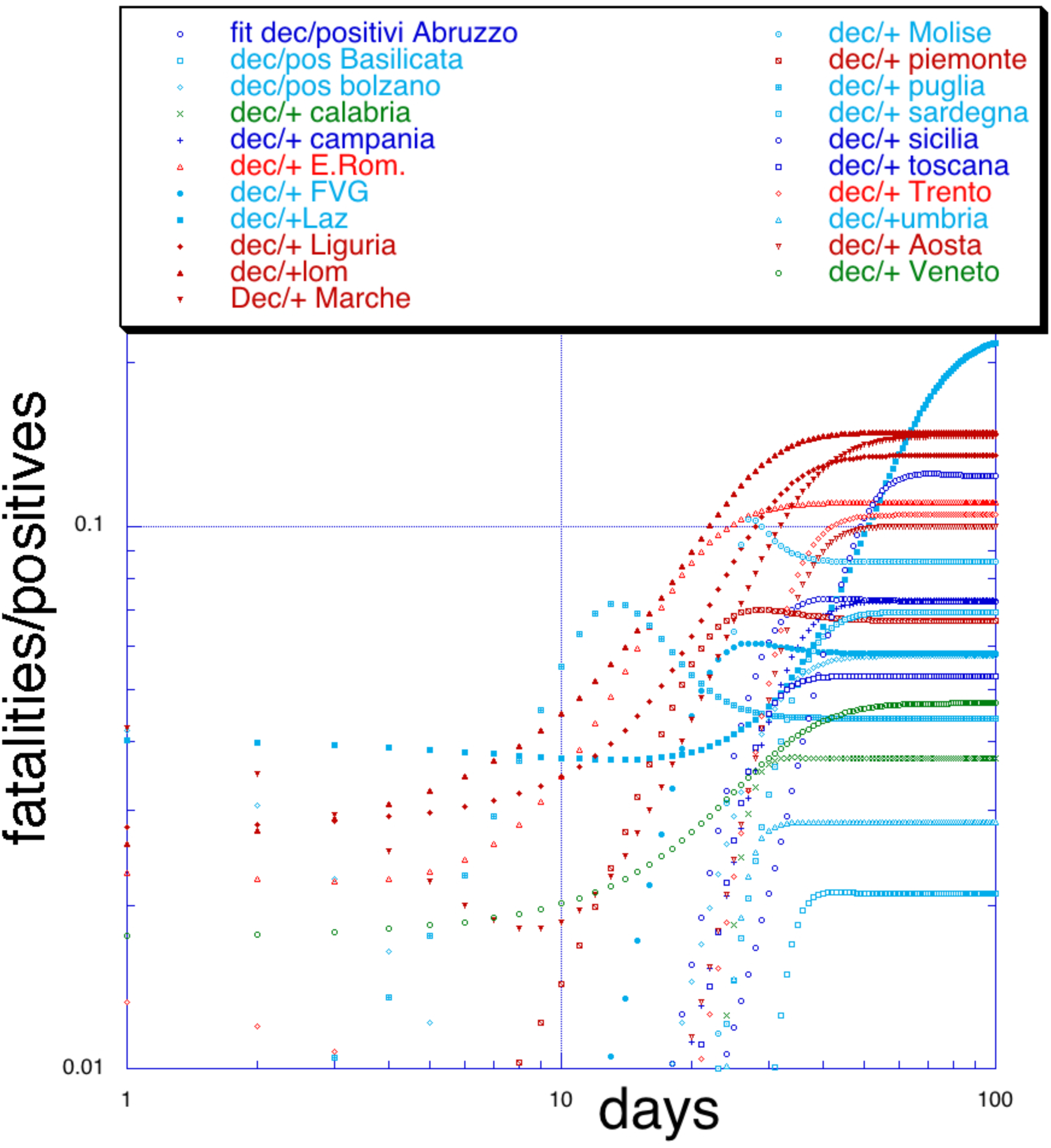
Fatalities/positives versus time, see the previous figures for the color codes. For comparison, the same ratio for Germany was 1.6% on April 5, 2020.

### Summary

We investigated in detail the spread of COVID-19 in each Italian region. The overall statistics shows that the spread is slowing down but not in some regions like Lombardy. The most negative feature is the statistical large number of fatalities as compared to the number of people tested positive to the virus. This is most probably due to hospital overcrowding and lack of enough tools like ventilators and sufficient personal protection equipment especially for the medical staff, which is in the front line. We cannot exclude that hospitals are a possible source of the infection, maybe the use of other public buildings transformed into temporary hospitals might help. Moving higher risk people, still negative to the virus, to lower density places like beach resorts might help. It is especially important in our opinion to put the different resources of the regions together to understand the spread more effectively. For instance regions with a lower probability of infection (less than 15%) might send some of their testing equipment, unused ventilators and maybe some medical personnel to higher risk regions on a temporary basis. Ventilators and personal protection equipment are the most needed tools. The country of Ferrari, Lamborghini and Ducati among others, as well as electronics and fashion might divert some industrial capabilities to fulfill the emergency in short times. The more we wait the more people dies.

## Data Availability

the data is available on request from the author

